# Changes in brain structure and function in a multisport cohort of retired female and male athletes, many years after suffering a concussion. The ICHIRF-BRAIN Study

**DOI:** 10.1101/2022.07.21.22277627

**Authors:** Michael Turner, Antonio Belli, Rudolph J. Castellani, Paul McCrory

**Affiliations:** The International Concussion and Head Injury Research Foundation (ICHIRF); The Institute of Sport, Exercise and Health, University College London, 170 Tottenham Court Road, London W1T 7HA, UK; Neurotrauma and Ophthalmology Research Group, Institute of Inflammation & Ageing, University of Birmingham, Birmingham B15 2TT, UK; and NIHR Surgical Reconstruction and Microbiology Research Centre, University Hospitals Birmingham NHS Foundation Trust, Birmingham, UK; and Marker Diagnostics UK Limited, the BioHub, Birmingham research park, Birmingham UK College of Medical and Dental Sciences, Institute of Cancer and Genomic Sciences, Centre for Computational Biology, University of Birmingham, B15 2TT, UK; Northwestern University, Feinberg School of Medicine, 420 East Superior Street, Chicago, Illinois 60611, USA

**Author notes:** **Correspondence to:** Dr Michael Turner, ICHIRF, 170 Tottenham Court Road, London W1T 7HA.

**Keywords:** Traumatic brain injury, concussion, sport, brain imaging, brain structure and function

## Abstract

Mild traumatic brain injury is widely regarded as a misnomer: it is globally a major cause of disability and is hypothesized as a potential causal factor in subsequent neurodegeneration. Commonly arising in sport, mounting evidence of varying degrees of cognitive impairment in retired athletes exposed to repeated concussions motivates close examination of its cumulative effects on the brain. Studying a cohort of 125 retired athletes with a mean of 11 reported concussions and 36 matched controls with none, here we evaluated whole-brain volumetric and subcortical morphological effects with Bayesian regression models and functional connectivity effects with network-based statistics. Estimates of potential cognitive impact were derived from meta-analytic functional mapping based on 13,459 imaging studies. Across the array of brain structural and functional effects identified, regions significantly lower in volume in the concussed group included, in order of greatest effect size, the middle frontal gyrus, hippocampus, supramarginal gyrus, temporal pole, and inferior frontal gyrus. Conversely, brain regions significantly larger within the athlete group included, in order of greatest effect size, the hippocampal and collateral sulcus, middle occipital gyrus, medial orbital gyrus, caudate nucleus, lateral orbital gyrus, and medial segment to the postcentral gyrus (all significant with 95% Bayesian credible interval). Subcortical morphology analysis corroborated these findings, revealing a significant, age-independent relationship between inward deformation of the hippocampus and the number of concussions sustained (corrected- *p*<0.0001). Functional connectivity analyses revealed a distinct brain network with significantly increased edge strength in the athlete cohort comprising 150 nodes and 400 edges (corrected-*p*=0.02), with the highest degree nodes including the pre-central and post-central gyri and right insula. The functional communities of the greatest eigenvector centralities corresponded to motor domains. Numerous edges of this network strengthened in athletes were significantly weakened with increasing bouts of concussion, which included disengagement of the frontal pole, superior frontal, and middle frontal gyri (*p*=0.04). Aligned to meta-analytic neuroimaging data, the observed changes suggest possible functional enhancement within the motor, sensory, coordination, balance, and visual processing domains in athletes, attenuated by concussive head injury with a negative impact on memory and language. That such changes are observed many years after retirement from impact sport suggests strong repetition effects and/or underpinning genetic selection factors. These findings suggest that engagement in sport may benefit the brain across numerous domains, but also highlights the potentially damaging effects of concussive head injury.

## Introduction

With a global incidence of 45-54 million individuals every year[1], mild traumatic brain injury (TBI) is a leading cause of disability worldwide, with some population-based epidemiological evidence that head injury is a relative risk for neurodegeneration and dementia, including Alzheimer’s disease[2]. A dose-dependent response is suggested, with the risk of dementia doubling following severe injury, and a 1.6-fold increase after mild TBI[2-6]with some developing symptoms more than 30 years after the initial traumatic insult[3]. Other studies have suggested little to no relationship between TBI and dementia or Alzheimer’s disease.[7-13]More than 43 million individuals suffer from some form of dementia globally, a figure that has more than doubled since 1990. It is currently the fifth leading cause of death[14]. Disentangling the underlying attributing factors to neurological decline – especially within the context of increasing incidence across an ageing population – is a clear research priority.

Between 15-20% of mild TBI prompting medical attention are attributable to sporting exposure[15, 16], with the incidence of sport-related TBI more than doubling since 1998[17]. Given that a significant proportion of underlying TBI is driven by a sporting aetiology, utilising such a cohort for models to study neurodegeneration becomes feasible. To that end, formally quantifying the impact of mild TBI resultant from concussive injury in sport is an area of significant interest, with evidence that concussion-exposed retired athletes display cognitive impairment[18, 19], depression[20], neuroimaging abnormalities[21-30] and/or potential changes to brain metabolism disproportionate with age[31].

Brain imaging is a key platform to investigate changes in structure and function resulting from TBI. Of note, most previous imaging TBI studies investigate exclusively men (or include a disproportionately small sample of women), despite ongoing evidence of a significant – and differential - impact on both sexes[32-35]. This notion indicates our current evidence base for TBI to be predominantly formulated upon men, underrepresenting women despite contemporaneous research illustrating women to suffer bouts of sporting-induced TBI both more frequently and more severely[32, 33]. Secondly, most neuroimaging studies focus on singular imaging modalities, studying either structure or function, leaving us in the dark to draw an inference as to the interplay between the two. There are no current models that satisfactorily combine structure and function in a single modality, so some parcellation is required to capture both neuroanatomy and function, however, this paper attempts to better understand the connection between these two entities.

Increasing bouts of concussive head impacts are hypothesized as a pathogenic factor in brain proteinopathies and progressive neurodegeneration[36-38], with increasing atrophy demonstrable after TBI that is linked to poorer clinical outcomes[39, 40]. Understanding the long-term consequences of exposure to head impacts and concussion in sport is essential, both for protecting athletes and identifying a cohort at increased risk on which preventative measures can be evaluated[41]. Athletes, therefore, represent a clearly defined group at potential risk of damage to brain structure and function from mild TBI, as well as a means to study models of long-term consequences of TBI.

In this study, we reveal the structural and functional brain changes of 125 retired male and female athletes with a history of previous concussion-induced TBI, contrasted to 36 matched controls with no history of concussion to determine the effect of sports participation, ageing, and/or prior injury on imaging outcomes.

## Materials and methods

### Ethical approval and trial registration

Staged ethical approval was obtained from St Mary’s University, Twickenham, Waldgrave Road, London TW1 4SX, UK initially on 1st June 2015 (no reference number was provided by the University Ethics Committee). Study approved.

27^th^ October 2015, as above, Reference SMEC.2015.16.53. Approved

12^th^ June 2017, as above, Reference SMEC.2016.17.115. Approved

26^th^ January 2018, as above, Reference SMEC.2017.18.051. Approved

In preparation for BREXIT, and before screening in Ireland, the whole study was re-submitted for ethical approval to the Beacon Hospital Research Ethics Committee (BHREC), The Beacon Hospital, Sandyford, Dublin D18 AK68, Ireland, and approved on 10^th^ October 2019. Reference BEA0130. An International, multicentre study into the long-term effects of concussion. The International Concussion and Head Injury Research Foundation (ICHIRF) Study. Fully Approved.

All participants provided informed consent in writing.

The trial is registered as ISRCTN 11312093.

### Study population

Participants were recruited by the International Concussion and Head Injury Research Foundation (ICHIRF) with the primary aim of investigating brain changes related to a previous history of concussion. All potential participants were pre-screened by completing a detailed online questionnaire regarding a concussion history, and physical and mental health status. 1,200 questionnaires were distributed initially and 787 were fully completed (51.9% male, 48.1% female). Of these, the 318 volunteers over the age of 50 were invited for screening in London (53.1% males, 46.9% females). Our inclusion criteria were individuals who completed the online screening and had previously competed in sport at a competitive level. Our exclusion criteria were any participants under the age of 50 years, those with a previous history of an intracranial bleed, those receiving any psychotropic medication, and any with a pre-existing medically diagnosed neurological disorder, including but not limited to Alzheimer’s disease, Parkinson’s disease, multiple sclerosis, and Motor Neuron Disease (Amyotrophic Lateral Sclerosis). Two discrete cohorts were recruited: i) a group of 125 retired athletes with exposures to concussion throughout their careers - 68 males and 57 females; mean age ± SEM 61.18 ± 0.82, and ii) age and gender-matched controls, with no previous episodes of concussion - 20 male and 16 females; mean age 62.17 ± 1.10 (Table 1 and Supplementary Figure 1). The nature of the sports played was deliberately varied – to enable a generalizable model of sport-induced concussion and mild TBI - and included boxing, cricket, cycling, equestrian, horse racing, martial arts, motorsports, rugby, skiing, and football (soccer).

**Table 1:** Cohort demographic and concussion exposure differences, with mean ± SEM and IQR illustrated. Only concussion status differed between the two groups. Age and gender did not differ significantly differ. Asterisks, † Two-tailed unpaired t-test assuming equal variance, ‡ Fisher’s exact test. Abbreviations; IQR. interquartile range; NSD. not significantly different; SEM, standard error of the mean.

| Variable | Concussed Athletes<br>(n=125) | Matched Controls<br>(n=36) | Statistical<br>Difference |
| --- | --- | --- | --- |
| Age (years) | $61.18 \pm 0.82$ (IQR 55-68) | $62.17 \pm 1.10$ (IQR 57-68) | NSD ($p = 0.54$) † |
| Gender | 68 male, 57 female | 20 male, 16 female | NSD ($p = 0.99$) ‡ |
| Number of<br>concussions<br>sustained | $10.68 \pm 1.10$ (IQR 4-13) | $0.00 \pm 0.00$ | $p < 0.0001$ † |

**Supplementary Figure 1:**
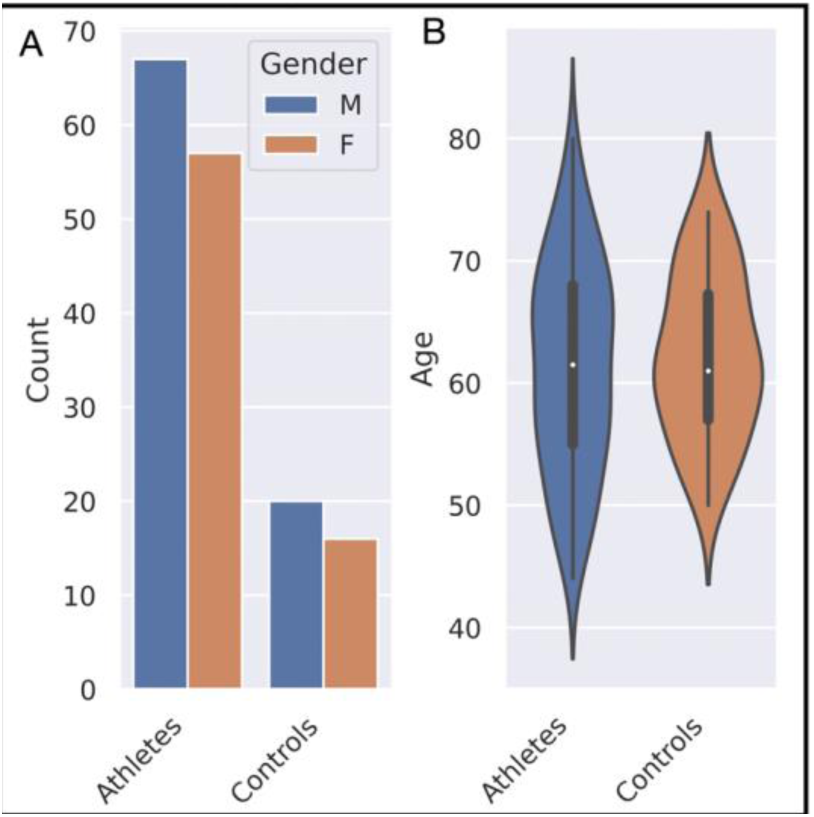
Cohort demographics and lifetime injury status. A) Count plot of sample sizes for athletes and controls, in addition to gender split, which did not significantly differ. B) Violin plot of age distribution between the groups, illustrating no significant difference. The groups are similar and balanced and no statistically significant difference was demonstrated between the two groups.

### Blinding

The statistical analysts were blinded to the allocation groups of participants in the assessment and analysis of all data.

### Acquisition of MRI data

Participants underwent brain imaging by magnetic resonance imaging on 3 Tesla Siemens scanners with a 32-channel head coil. The acquired sequences that were used in our analysis were the following: 1) T1-weighted structural image: a high-resolution 3D image was acquired using the Magnetization Prepared Rapid Gradient Echo (MPRAGE) sequence with the following parameters: Repetition Time (TR) 2000ms; Echo Time (TE) 2.98ms; Inversion Time (TI) 900ms; flip angle 9 degrees; Field of View (FOV) 144 mm x 240 mm x 256 mm; voxel size 1.2mm x 1mm x 1mm, of acquisition time 8 minutes and 2 seconds. 2) Resting-state functional MRI (rsfMRI): T2*-weighted images were acquired during which participants rested with their eyes shut using a Gradient-Echo Echo-Planar-Imaging (EPI) sequence. A total of 165 volumes were acquired, each containing 74 axial slices, slice thickness 3mm with an interslice gap of 10%, providing whole-brain coverage from vertex to cerebellum; TR 2500ms; TE 25ms; flip angle 90 degrees; FOV 219mm x 219mm; voxel size 2.9mm x 2.9mm x 3.30mm; acquisition time of 6 minutes 52 seconds.

### Imaging analysis pipeline

The curation of multi-modal structural and functional brain imaging permitted an in-depth investigation of the brain differences across the cohort of interest that included structural evaluation at the cortical and subcortical level, in addition to functional brain connectivity. The schematic for our overall approach is demonstrated in Supplementary Figure 2. Before any image pre-processing, all sequences were carefully reviewed manually for signal and image artefacts that may have otherwise confounded findings. Similarly, all scans were further re-reviewed at every pre-processing stage to ensure no fault which may confound findings.

**Supplementary Figure 2:**
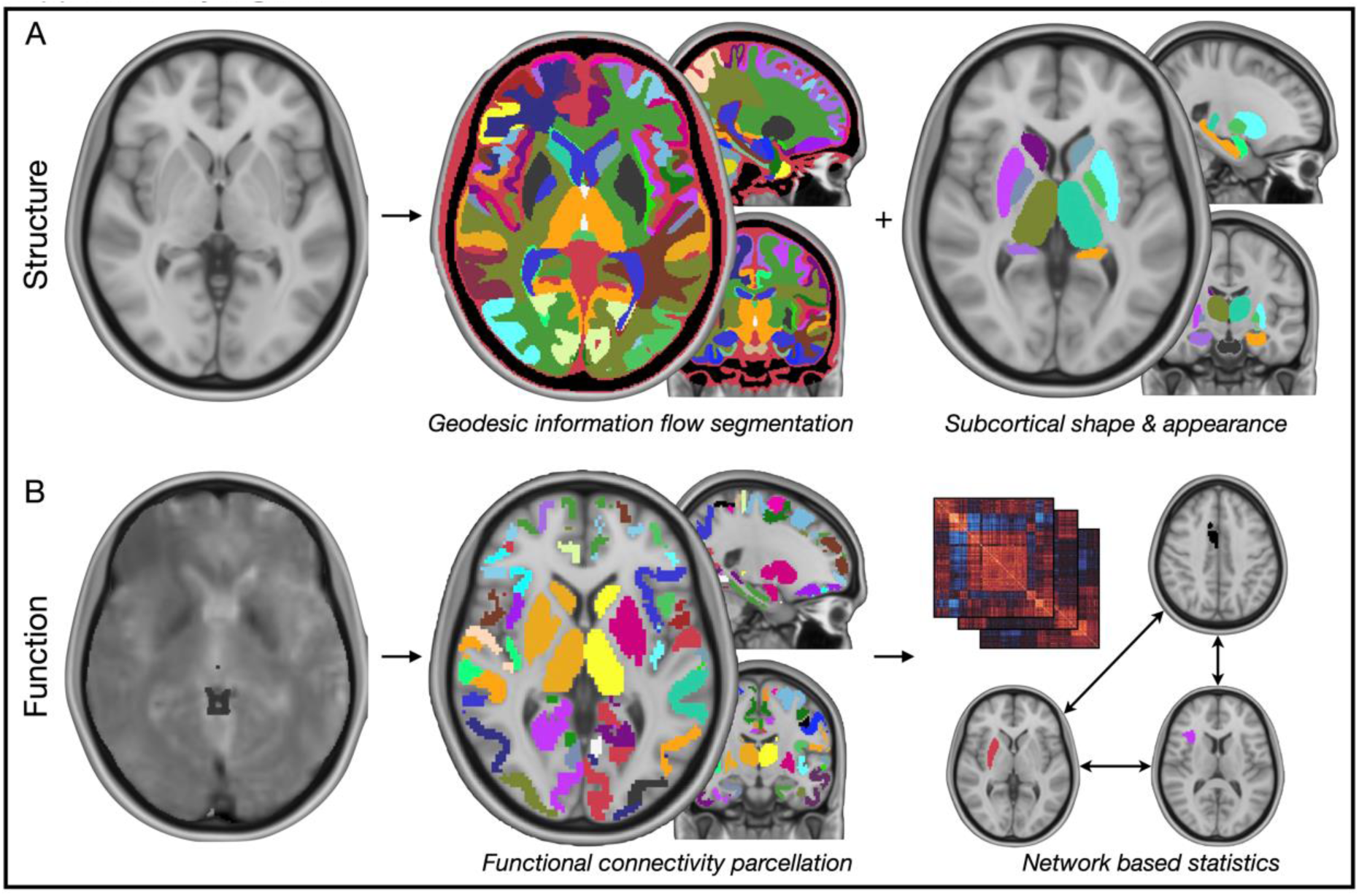
Imaging processing pipeline. A) Structural analyses were undertaken in two distinct domains: i) whole brain segmentation and parcellation by geodesic information flows for analysis of grey matter volumes and ii) subcortical shape and appearance with FSL-FIRST. B) Functional imaging data were partitioned into a parcellation scheme of 346 regions, derived connectivity matrices across the cohort, and used this data for network-based statistics to assess for significant functional brain networks related to cohort differences and examined how this might be affected by concussion.

### Structural neuroimaging

We investigated structural neuroanatomical differences between the concussed athletes and controls in two distinct approaches: i) robust whole-brain parcellation for grey matter volumetry[42] and ii) morphological assessment of the subcortex[43]. The rationale for this was to ascertain differences not merely in underlying volumes of brain structures contingent on the group or concussion exposure, but additionally, reveal underlying changes in shape and appearance of brain structures.

Image processing was in line with the UK Biobank 10,000 imaging dataset [44]

#### Whole-brain parcellation

was pre-processed using geodesic information flows and NiftySeg (http://niftyweb.cs.ucl.ac.uk/)[42], a previously published pipeline encompassing spatially-variant graphs applied to probabilistic brain segmentation regardless of morphological variability. For each participant, this approach generated a parcellation of 162 neuroanatomical regions, from which we would utilise the total cubic millimetre volume of regions in subsequent multivariate models. Our priority in this approach was to evaluate volumetric differences in grey matter structures across the two groups and, with no specific prior hypothesis for a laterality effect, we extracted grey matter volumes from this data and pooled left and right regions to maintain degrees of freedom and reduce the risk of rank deficient variable orthogonality.

This modelling equation was chosen to reflect the cohort changes against all brain changes, thus ensuring that any regional brain changes are automatically conditioned/corrected to the other changes.

#### Bayesian inference of grey matter volumes

was undertaken with R packages ‘UPG’[45] for logistic regression between cohorts, a framework that can handle imbalanced data where sampling efficiency is maximised through Markov Chain Monte Carlo (MCMC) boosting, and ‘bayesreg’[46] for further regression tasks in modelling concussion. Input variables were the grey matter volumes, participant age, gender, and an intercept term. Raw regional volumes were non-normally distributed and therefore log-transformed to normality, with all features z-scored and scaled to ensure meaningful comparisons across beta coefficients. We computed 1,000,000 draws with a burn-in of 50,000, utilizing standard defaults for a prior variance of the coefficients as 1, prior shape and rate for the working parameter delta as 0.5, and prior variance for the intercept as 99. From this logit model, we extracted the posterior estimate and 95% credible interval (CI) with our criterion for a significant finding one where the beta coefficient CI did not cross zero. We considered three principal models to fit: i) a logit task of brain differences underpinning differences between the concussed athletes and matched controls; ii) this same logit but supplemented with the additional predictor of concussion, to ascertain what (if any) brain difference remained after controlling for this factor, and iii) a Poisson regression task to model the number of concussions from the interaction factor between gender and brain volumes.

#### Subcortical morphology

was pre-processed and analysed were performed using FSL-FIRST 5.0, a Bayesian modelling toolkit developed for segmentation, shape, and volumetric analysis of the subcortex[43]. This package firstly skull-strips with the FSL brain extraction tool (BET), following which performs segmentation of each patient structural scan into 15 subcortical structures: bilateral nucleus accumbens, amygdala, caudate, hippocampus, pallidum, putamen, thalamus, as well as the brainstem. Segmented structures are then registered to the Montreal Neurological Institute (MNI) 152 1mm template. The morphometry (shape deformations) of the subcortex were statistically tested with non-parametric inference with FSL-RANDOMISE, wherein we evaluated for differences between the cohorts with nuisance covariates of age, gender, and scanner site. A posthoc correction was employed by threshold-free-cluster-enhancement (TFCE) with our criterion for statistical significance a corrected *p* < 0.05.

### Functional neuroimaging

#### Pre-processing of resting-state functional magnetic resonance imaging (fMRI)

was undertaken using the FMRI Expert Analysis Tool (FEAT), version 6.00, within FSL[47]. The first 4 volumes of each 4-dimensional fMRI time series were always discarded to allow for signal stabilization. The following pre-statistical processing steps were applied: Motion Correction with FMRIB Linear Image Registration Tool (MCFLIRT) (7 degrees of freedom); slice-timing correction using Fourier-space time-series phase-shifting; brain extraction (BET); spatial smoothing using a Gaussian kernel of full-width-half-maximum (FWHM) 5mm; grand-mean intensity normalization of the entire 4-dimensional dataset by a single multiplicative factor; high pass temporal filtering (Gaussian-weighted least-squares straight-line fitting, with sigma = 50.0s) and registration to high resolution structural and standard space images using the FMRIB Linear Image Registration Tool (FLIRT)[48].

#### Whole-brain parcellation

of fMRI data was undertaken using a well-utilised functional scheme[49], further supplemented by the inclusion of the FSL-FIRST 15 regions of the subcortex to form a whole-brain parcellation of 346 unique regions. Functional connectivity adjacency matrices were generated by extraction and cross-correlation of time-sequence blood oxygen level-dependent (BOLD) signal. For each subject, these approaches produced adjacency matrices of 59685 functional edges covering all pairwise links between all brain regions. Connectivity matrices were r to z transformed.

#### Network-based statistics of functional connectivity

were undertaken using the networks-based statistics (NBS) connectome toolbox (v1.2[50, 51]). NBS is a non-parametric statistical method that corrects for multiple comparisons and controls for family-wise error rate (FWE). The NBS is a graph analogue of cluster-based statistical methods used in mass-univariate testing on all voxels in an image and produces clusters in topological space, as opposed to physical space. The NBS relies on permutation testing (Freedman & Lane method[52]) to determine significance within the general linear model, which includes regression of nuisance predictors, permuting resulting residuals and subsequently adding permuted residuals back to nuisance signal to give a realization of data under the null hypothesis. This approach recognises that permuting raw data is not desirable as it may engender some variability explained by nuisance predictors. Rather, it is the error terms that can be permuted and estimated under the null hypothesis as a part of the data not explained by the nuisance regressors; that is, the residuals[53]. The method permits the derivation of FWER-corrected *p* values using permutation testing when investigating brain networks[54]. Our statistical test was a comparison of the athletes with a history of concussion, against the controls. In posthoc analyses, we evaluated network differences as a function of the number of concussions an athlete had experienced. Statistical analysis included nuisance covariates of age and gender (all functional imaging was undertaken at the same centre, so need not be included as a covariate), with 50000 permutations, a minimum t-statistic threshold of 3.1, and our criterion for statistical significance was FWER *p* < 0.05.

#### Post-hoc analyses of functional connectivity networks

were undertaken to further interrogate the results identified. To do so, we extracted t-statistic weighted adjacency matrices for visual network analysis and derivation of centrality measures, using graph-tool[55]. Specifically, we quantified the weighted eigenvector centrality of graphical models contingent on the edge-weight of a given model fit (weighted to cohort and number of concussions), including a statistical comparison between well-described functional communities included within the parcellation scheme (including default mode, somatomotor, auditory, visual, *et cetera*)[49]. We also quantified the proportionate degree count of functional communities within significant networks, defined as its number of significant links normalised by the number of nodes within that functional community, to prevent over-representing a community merely because of its high nodal count within the parcellation scheme.

#### Cognitive implications from structural brain changes

were evaluated using meta-analytic functional maps derived from natural language processing of published imaging studies. This was achieved with NeuroQuery image search matching[56] in a framework similar to other published approaches[57]. Passing each identified significant ROI from Bayesian structural imaging models to the NeuroQuery repository of 13,459 studies encompassing 5,144 activation pattern terms, we retrieved the closest matching 10 topic terms. We filtered terms to exclude those that were either anatomical, experimental, vague, or disease descriptors, utilizing the Terminologia Anatomica dictionary to automatically filter out anatomical terms[58]. These terms were then weighted by the beta coefficients from each ROI, yielding a list of terms for each ROI, each term with a matching score, from which we generated a corpus with detailed information on the term frequency favouring both increased function in the concussed athlete group, and those of putatively decreased function.

### Data and code availability

Raw participant data is not suitable for dissemination under the consent framework. All software used is freely available via original referenced sources in the methods and we make model fits that contain no identifiable participant data freely available for use within the supplementary materials.

## Results

### Participant demographics, exposures, and concussion data

Age and gender did not significantly differ between the groups (Table 1 and Supplementary Figure 1). Only the athlete group had experienced previous episodes of concussion, with a mean number of concussive episodes of 10.68 ± 1.10, and no episodes of a concussion sustained across the matched controls (*p* < 0.0001).

### Structural brain differences

#### *Neuroradiology* reporting of imaging

across the cohort revealed 12 concussion subjects with clinical neuroradiological evidence of previous TBI (0 in controls) and 17 with chronic microangiopathic white matter change (2 in controls).

#### Bayesian cohort models of grey matter volumetry

by geodesic information flows segmentation and parcellation revealed many regions that significantly differed between the athletes with prior concussion, compared to the matched controls (Figure 1 <u>- below</u>). Brain regions significantly lower in volume in the athlete group included, in order of greatest effect size, the middle frontal gyrus, hippocampus, supramarginal gyrus, temporal pole, and inferior frontal gyrus. Conversely, brain regions significantly larger within the athlete group included, in order of greatest effect size, the hippocampal and collateral sulcus, middle occipital gyrus, medial orbital gyrus, caudate nucleus, lateral orbital gyrus, and medial segment to the postcentral gyrus. The full multivariate model posterior estimates are provided in Supplementary Table 1. Importantly, a re-analysis of this model - but with the inclusion of the number of concussions as an additional model covariate - rendered all brain differences non-significantly different between the two groups (<u>Supplementary Table 2</u>).

**Figure 1:**
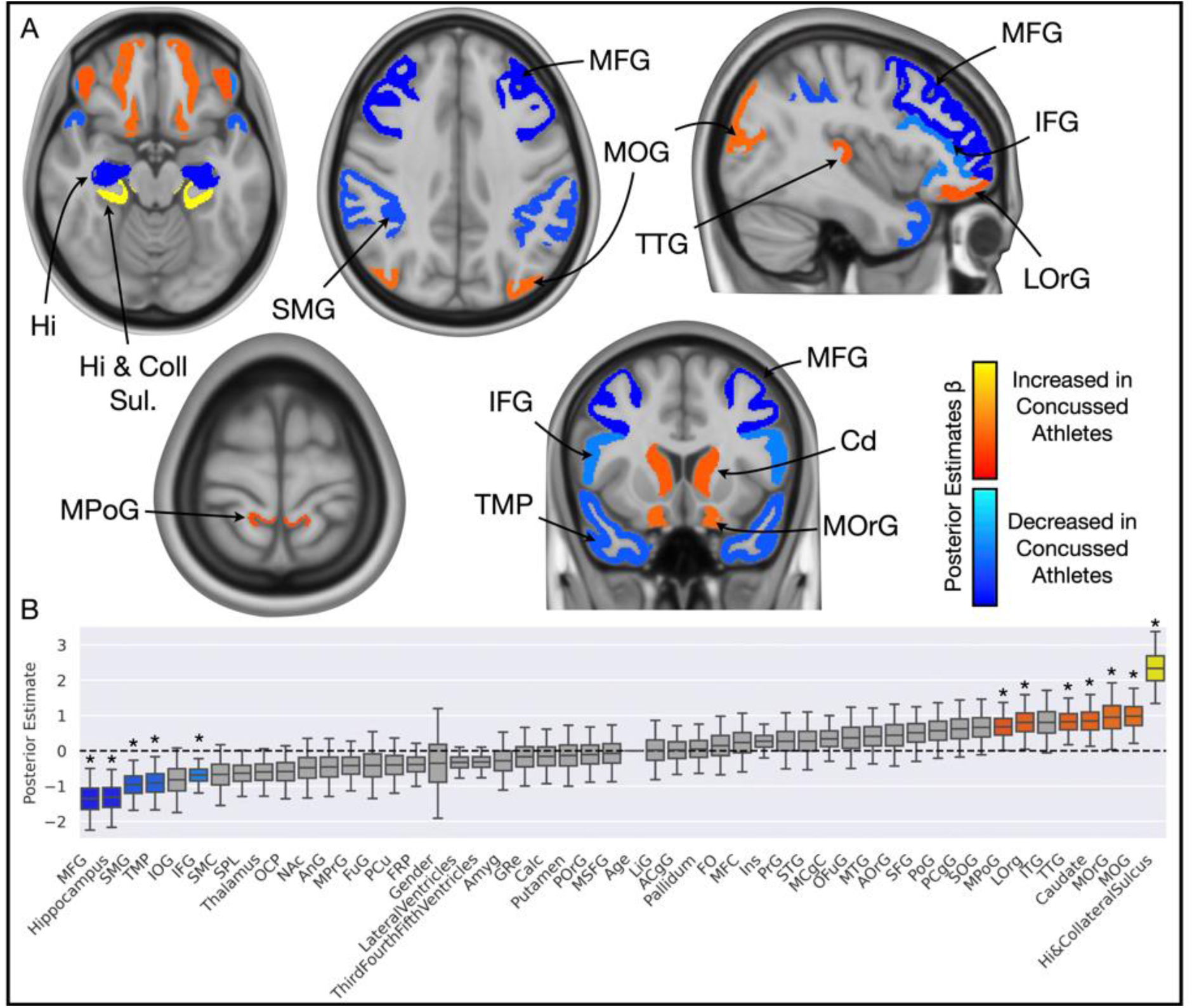
Grey matter volumetric differences across the cohort. A) Brain slices and B) boxplot illustrate colour-coded posterior estimates of betas from Bayesian logit, wherein red-orange-yellow regions are those found to increase in volume in the athlete group, compared to the controls; reciprocally, blue and light blue regions are those found to decrease in volume in the athlete group, compared to the controls. Only significant regions are shown on the brain images, and the colour is also aligned to the boxplot. Abbreviations: Amyg., amygdala; ACgG, anterior cingulate gyrus; AOrG, anterior orbital gyrus; AnG, angular gyrus; Calc, calcarine cortex; FO, frontal operculum; FRP, frontal pole; FuG, occipital fusiform gyrus; GRe, gyrus rectus; Hi, hippocampal [sulcus]; Ins, insula; IFG, inferior frontal gyrus; IOG, inferior occipital gyrus; ITG, inferior temporal gyrus; LiG, lingual gyrus; LOrg, lateral orbital gyrus; MCgG, middle cingulate gyrus; MFC, medial frontal cortex; MFG, middle frontal gyrus; MOG, middle occipital gyrus; MOrG, medial orbital gyrus; MPoG, postcentral gyrus medial segment; MPrG, precentral gyrus medial segment; MSFG, superior frontal gyrus medial segment; MTG, middle temporal gyrus; NAc, nucleus accumbens; OCP, occipital pole; OFuG, occipital fusiform gyrus; PCgG, posterior cingulate gyrus; PCu, precuneus; PoG, postcentral gyrus; POrG, posterior orbital gyrus; PrG, precentral gyrus; SFG, superior frontal gyrus; SMC, supplementary motor cortex; SMG, supramarginal gyrus; SOG, superior occipital gyrus; SPL, superior parietal lobule; STG, superior temporal gyrus; TMP, temporal pole; TTG, transverse temporal gyrus.

#### Bayesian gender-concussion regression models of grey matter volumetry

revealed a differential pattern of brain volume differences related to concussion, depending on participant gender, with a pseudo-r2 of 0.58. In this cohort, the male gender favoured an overall higher number of concussions reported. In order of greatest effect size, in males, an increasing number of concussions was associated with decreased volume to the superior parietal lobule, precuneus, fusiform gyrus, angular gyrus, occipital pole, middle frontal gyrus, middle occipital gyrus, occipital fusiform gyrus, and insula, with an increase in the volume of the third and fourth ventricles. However, in females, an increasing number of concussions was associated with decreased volume of the postcentral gyrus, nucleus accumbens, supramarginal gyrus, caudate nucleus, inferior occipital gyrus, and middle orbital gyrus, with an increase in the volume of the lateral ventricles. The interaction between female gender and increasing age tended towards a higher number of concussions, though 95% credible intervals marginally crossed zero. Age alone was not a predictor of the number of concussions sustained. The full multivariate model posterior estimates are provided in <u>Supplementary Table 3.</u>

#### Subcortical shapes and appearance analyses

identified several brain regions that significantly differed in morphology between the two groups (Figure 2 - below). The bilateral hippocampus (left, corr-*p* < 0.0001; right, corr-*p* = 0.02), left thalamus (corr-*p* = 0.007) and left amygdala (corr-*p* = 0.01) all favoured significant inward morphological deformation in the concussed athlete group, compared to the controls, whilst the left caudate nucleus favoured outward morphological deformation in the athletes, compared to controls (corr-*p* = 0.02). We also evaluated if the number of concussion bouts related to morphological differences, which identified a significant relationship between the number of concussions ascertained and inward deformation to the hippocampus bilaterally (left, corr-*p* < 0.0001; right, corr-*p* = 0.004) and the left amygdala (corr-*p* = 0.0002). We further investigated these areas for a function of age, which identified a significant relationship between increasing age and inward deformation at the bilateral hippocampus (left, corr-*p* < 0.0001; right, corr-*p* = 0.02), left thalamus (corr-*p* = 0.02) and amygdala (corr-*p* = 0.03) but observed the significant margins affected largely different aspects of the regional surface. This is further detailed in Supplementary Figure 3 with companion case examples. The athletes with a history of concussion illustrated a significant linear effect for inward deformation of hippocampus vertex surfaces, bilaterally, compared to controls (*p* < 0.0001). Inward hippocampal deformation was significantly negatively correlated to increasing age for both athletes and controls (r – 0.12, *p* < 0.0001), but the gradient of linear effect non-significantly differed between the two groups.

**Figure 2:**
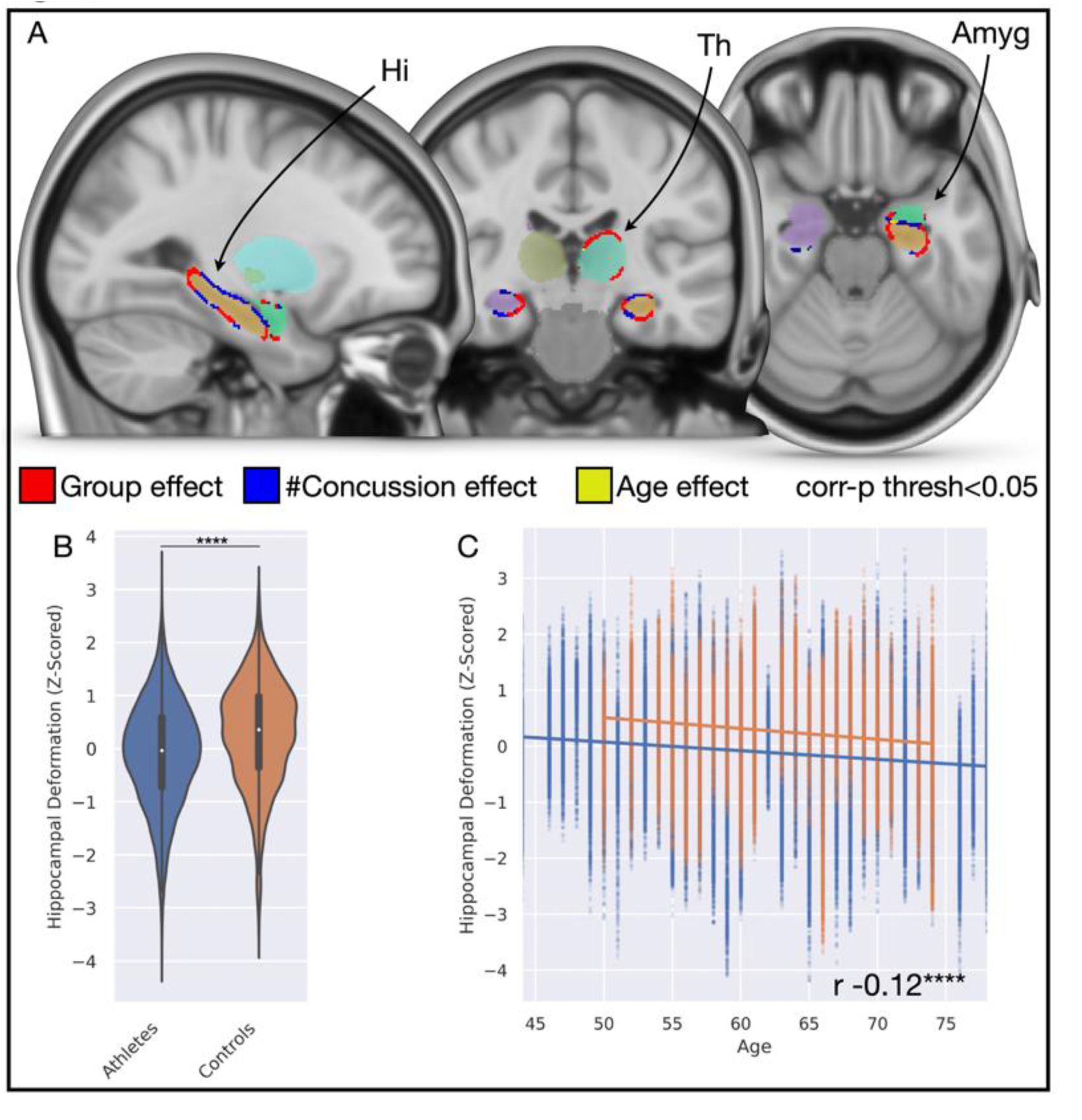
Subcortical shape and appearance differences across the cohort. A) Brain slices illustrate significant differences in morphology of the hippocampus bilaterally, the left thalamus, and left amygdala with associations to group (red), the number of concussions (blue), and age (yellow). B) Violin plot illustrates bilateral hippocampal shape deformation significantly favours *inward* deformation in the athlete group, compared to controls. C) Scatterplot with linear regression illustrates that hippocampal shape significantly tends to inward deformation with increasing age in both groups, but that the gradient decline is not significantly different between groups, indicative that findings are not merely driven by a function of age. Abbreviations: Amyg, amygdala; Hi, hippocampus; Th, thalamus.

**Supplementary Figure 3:**
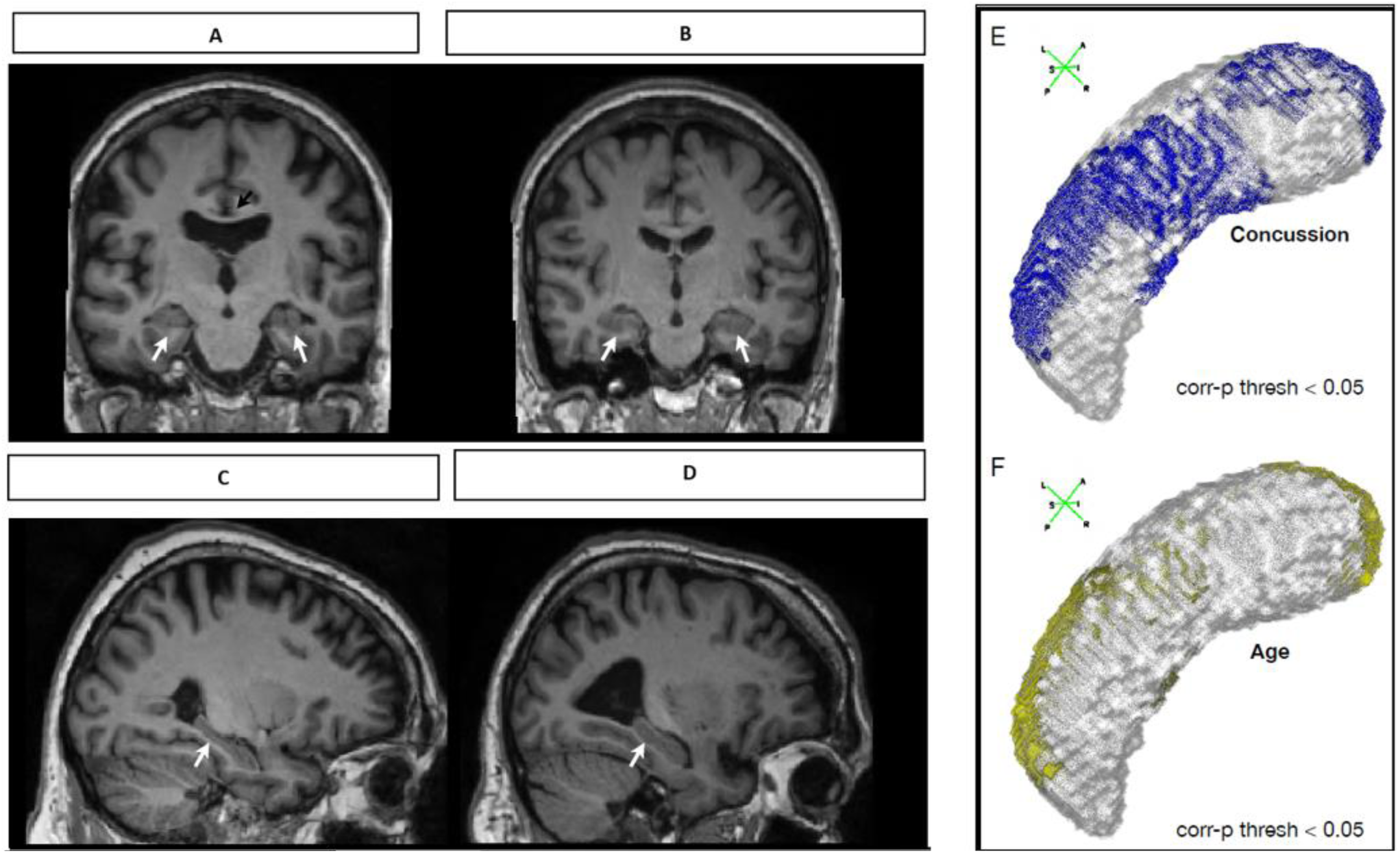
Case example images across the cohort. A) Coronal T1-weighted MRI of a male aged 60-66 within the concussed athlete group, positioned at the level of the mid hippocampus (white arrows), a cavum septum pellucidum is also noted (black arrow). B) Paired T1-weighted MRI of a man aged 60-66 from within the control group, positioned at the level of the mid hippocampus (white arrows). The hippocampi of participant A) are appreciably smaller than in B), C) Sagittal T1-weighted MRI of a man aged 70-75 within the concussed athlete group, who over their career recorded a total of over 20-25 episodes of concussion, positioned at the level of the hippocampus (white arrow). D) Paired sagittal T1-weighted MRI of a man aged 70-75 within the concussed athlete group, who over their career recorded a total of 1-5 episodes of concussion, positioned at the level of the hippocampus (white arrow). Despite evidence of more marked generalized atrophy within participant D), the midpoint of the hippocampus in participant C) - the individual with far more episodes of concussion documented - is appreciably thinner. E) 3D vertex plot of the significant vertices of the hippocampus, wherein blue foci show significant inward shape deformation related to increased bouts of concussion. F) Paired 3D vertex plot of the significant vertices of the hippocampus, wherein yellow foci show significant inward shape deformation related to increased age. Whilst there are similarities between both E-F), the effect of concussion appreciable affects more the hippocampal mid-portion, corroborating the example participant findings in panel C-D).

### Functional brain differences

#### Changes in the functional brain networks in athletes

Network-based statistics comparing brain network differences between the athletes and controls identified a significant functional network – the strength of which was *increased –* within the athlete cohort. At a *t-statistic* threshold of > 3.1, this comprised 150 nodes and 400 edges (corr-*p* = 0.02) which, at a *t-statistic* threshold of > 3.5, maintained 71 nodes and 121 edges (corr-*p* = 0.02) (Figure 3 <u>- below</u>). This network encompassed a breadth of brain regions, including nodes such as the pre-and post-central gyri, insula, middle frontal gyrus, cingulate cortex, central operculum, superior frontal gyrus, and precuneus. Pooled connectivity across the entire network was significantly greater within the athlete group (*p* < 0.0001). A review of the degree of centrality of these regions illustrated a predominance of significant edges to connect the precentral gyrus, insula, postcentral gyrus, and central opercular cortex. Quantifying the weighted eigenvector centrality of these nodes revealed that, when organised by functional community, the somatomotor, visual, and auditory nodes (in descending order) had significantly greater eigenvector centrality, compared to the remainder (ANOVA *p* < 0.0001). Reviewing the proportionate degree counts of these communities confirmed the greatest degree centralities were for auditory and somatomotor networks (in descending order). No significant difference related to gender with group allocation was observed.

**Figure 3:**
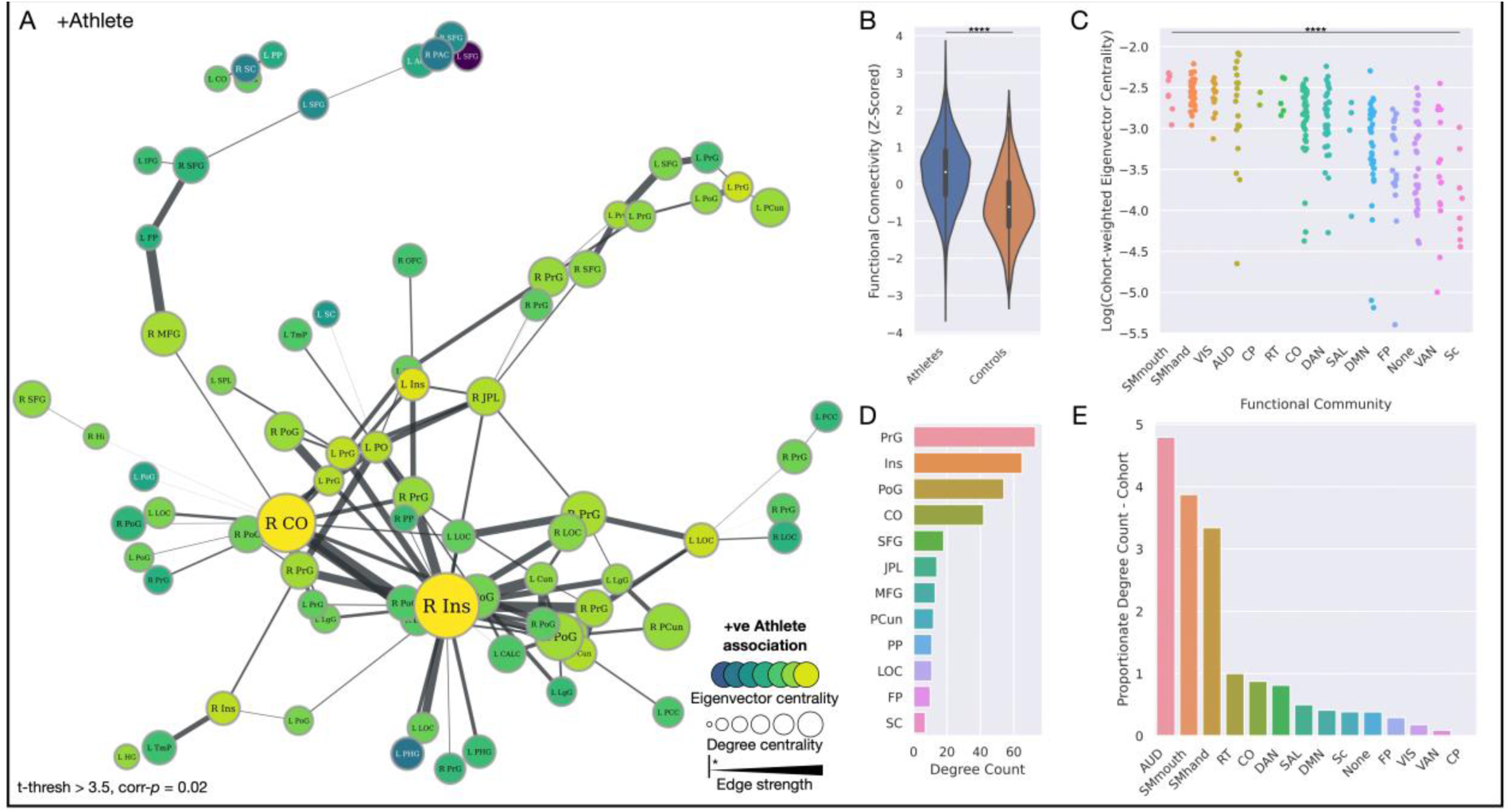
A strengthened functional brain network in athletes. A) Network plot illustrating the significant functional network of increased strength in athletes, with the size of nodes proportionate to their degree centrality, and colour proportionate to their group-weighted eigenvector centrality. Edge width is proportionate to the t-statistic of the group difference edge strength. All edges shown are significant. B) Violin plot illustrating functional connectivity of this network to be significantly greater in the athlete group. C) Strip plot illustrating a significant difference in eigenvector centrality of brain regions across all functional communities, weighted by t-statistic of the significant athlete network, ordered by decreasing values of centrality. D) Bar plot depicting degree counts of the most prominent nodes in the network. E) Bar plot illustrating degree counts of functional communities, proportionate to the number of nodes that feature in each community. *Community abbreviations*: AUD, auditory; CO, cingulo-opercular; CP, cingulo-parietal; DAN, dorsal attention; DMN, default mode network; FP, fronto-parietal; RT, retrosplenial temporal; SAL, salience; Sc, subcortical; SMhand, somatomotor hand; SMmouth, somatomotor mouth; VAN, ventral attention; VIS, visual. *Node abbreviations*: ACC, anterior cingulate cortex; CALC, intracalcarine cortex; CO, central opercular cortex; Cun, cuneus; FP, frontal pole; Hi, hippocampus; HG, Heschel’s gyrus; IFG, inferior frontal gyrus; Ins, insula cortex; JPL, juxtapositional lobule; L, left; LgG, lingual gyrus; LOC, lateral occipital cortex; MFG, middle frontal gyrus; OFC, orbitofrontal cortex; PAC, paracingulate gyrus; PCC, posterior cingulate cortex; PCun, precuneus cortex; PHG, parahippocampal gyrus; PO, parietal operculum; PoG, postcentral gyrus; PP, planum polare; PrG, precentral gyrus; R right; SC, subcallosal cortex; SFG, superior frontal gyrus; SPL, superior parietal lobule; TmP, temporal pole.

#### A functional brain network in athletes, inversely related to episodes of concussion

Having identified a functional brain network strengthened within the athlete group, we evaluated if this network strength was related to the number of episodes of concussion. Examining the athlete cohort only – since this was the only group with episodes of concussion – illustrated several of these network links to be inversely related to the number of episodes of concussion – i.e., which decreased in strength as the number of concussive episodes increased (ANOVA *p* = 0.04). Specifically, this included links between the left and right middle frontal gyrus, the left and right frontal pole, the left and right superior frontal gyrus, and the links between the middle frontal gyrus, frontal pole, and superior frontal gyrus. In addition, edge strength between the cuneus and superior frontal gyrus, as well as precentral gyrus and insula were inversely related to the number of concussions. Investigating per gender, this significant decrease in connection strength related to increasing counts of concussion was identified only in the male sample (*p* = 0.01), with no significant difference apparent in the females (*p* = 0.78). Figure 4 <u>- below</u>.

**Figure 4:**
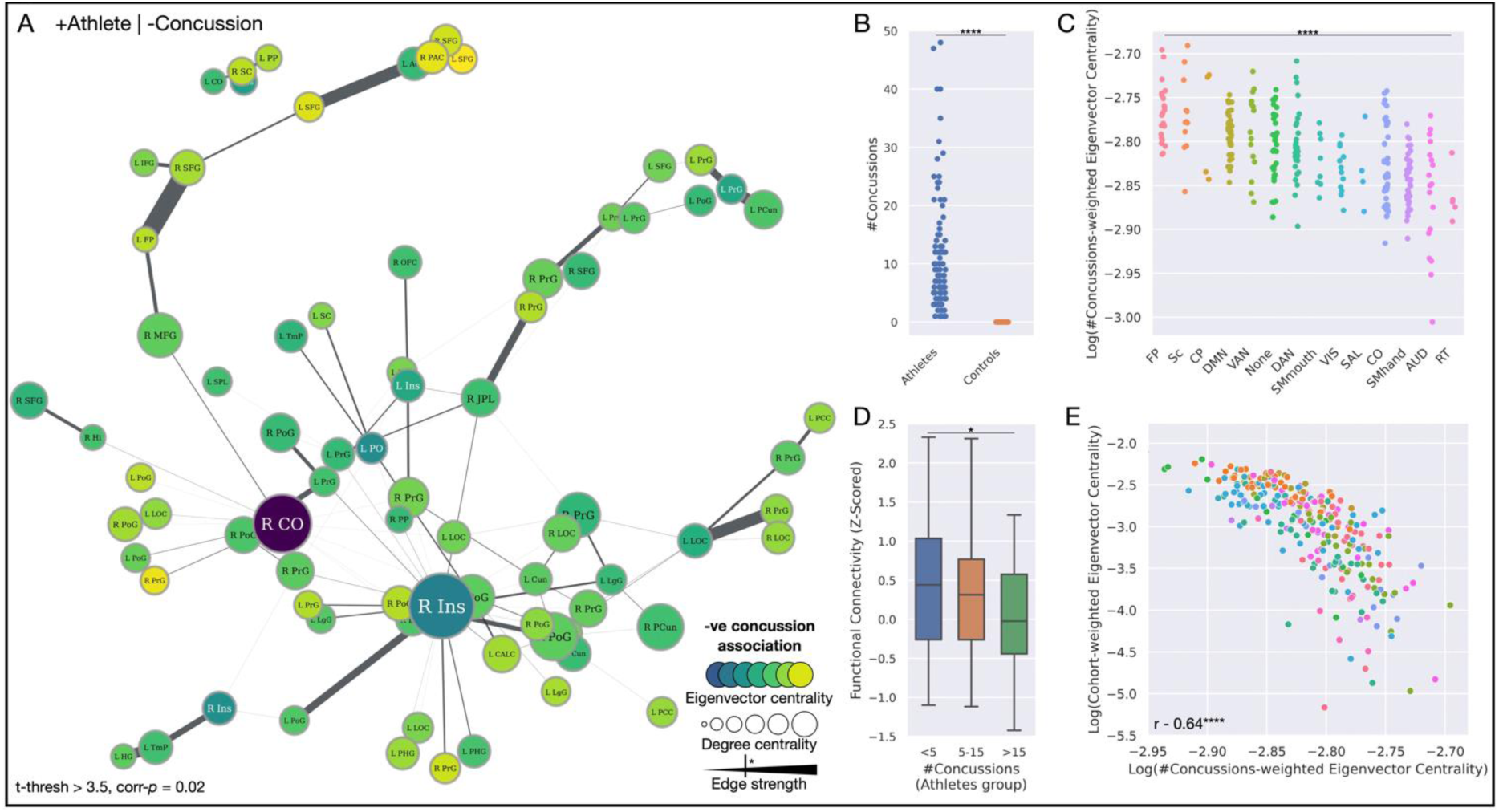
A strengthened functional brain network in athletes but weakened with increasing episodes of concussion. A) Network plot illustrating the significant functional brain network, with the size of nodes proportionate to their degree centrality, and colour proportionate to their concussion-weighted eigenvector centrality. Edge width is proportionate to the t-statistic of the edge strength related to a decreasing number of concussions (i.e., edge strength is inversely related to the number of concussions). Significant edges are depicted by the key. B) Strip plot illustrating the significant spread of a number of concussions exclusively across the athlete cohort. C) Strip plot illustrating a significant difference in eigenvector centrality of brain regions across all functional communities, weighted by t-statistic of its inverse relation to the number of concussion episodes, ordered by decreasing values of centrality. D) Bar plot illustrating that the strength of the edges significantly decreases with a rise in concussion. E) Scatterplot illustrating the significant inverse relationship between the centrality of brain regions according to strength in the athlete cohort, compared to their relationship to concussion. *Community abbreviations*: AUD, auditory; CO, cingulo-opercular; CP, cingulo-parietal; DAN, dorsal attention; DMN, default mode network; FP, fronto-parietal; RT, retrosplenial temporal; SAL, salience; Sc, subcortical; SMhand, somatomotor hand; SMmouth, somatomotor mouth; VAN, ventral attention; VIS, visual. *Node abbreviations*: ACC, anterior cingulate cortex; CALC, intracalcarine cortex; CO, central opercular cortex; Cun, cuneus; FP, frontal pole; Hi, hippocampus; HG, Heschel’s gyrus; IFG, inferior frontal gyrus; Ins, insula cortex; JPL, juxtapositional lobule; L, left; LgG, lingual gyrus; LOC, lateral occipital cortex; MFG, middle frontal gyrus; OFC, orbitofrontal cortex; PAC, paracingulate gyrus; PCC, posterior cingulate cortex; PCun, precuneus cortex; PHG, parahippocampal gyrus; PO, parietal operculum; PoG, postcentral gyrus; PP, planum polare; PrG, precentral gyrus; R right; SC, subcallosal cortex; SFG, superior frontal gyrus; SPL, superior parietal lobule; TmP, temporal pole.

### Meta-analysis of cognitive function aligned to brain structural differences

Using the posterior estimates of these significantly different brain regions, we computed a weighted functional corpus for both regions increased and decreased in volume in the athlete cohort with a history of concussion to identify putative cognitive changes. A range of terms spanning motor, sensory, perception, balance, coordination, visual processing, and coordination demonstrated increased weighting with putatively functional enhancement within the concussed athlete cohort. Conversely, however, functional attenuation within the concussed athlete cohort predominated around memory, learning, recall and natural language (Figure 5 <u>- below</u>).

**Figure 5:**
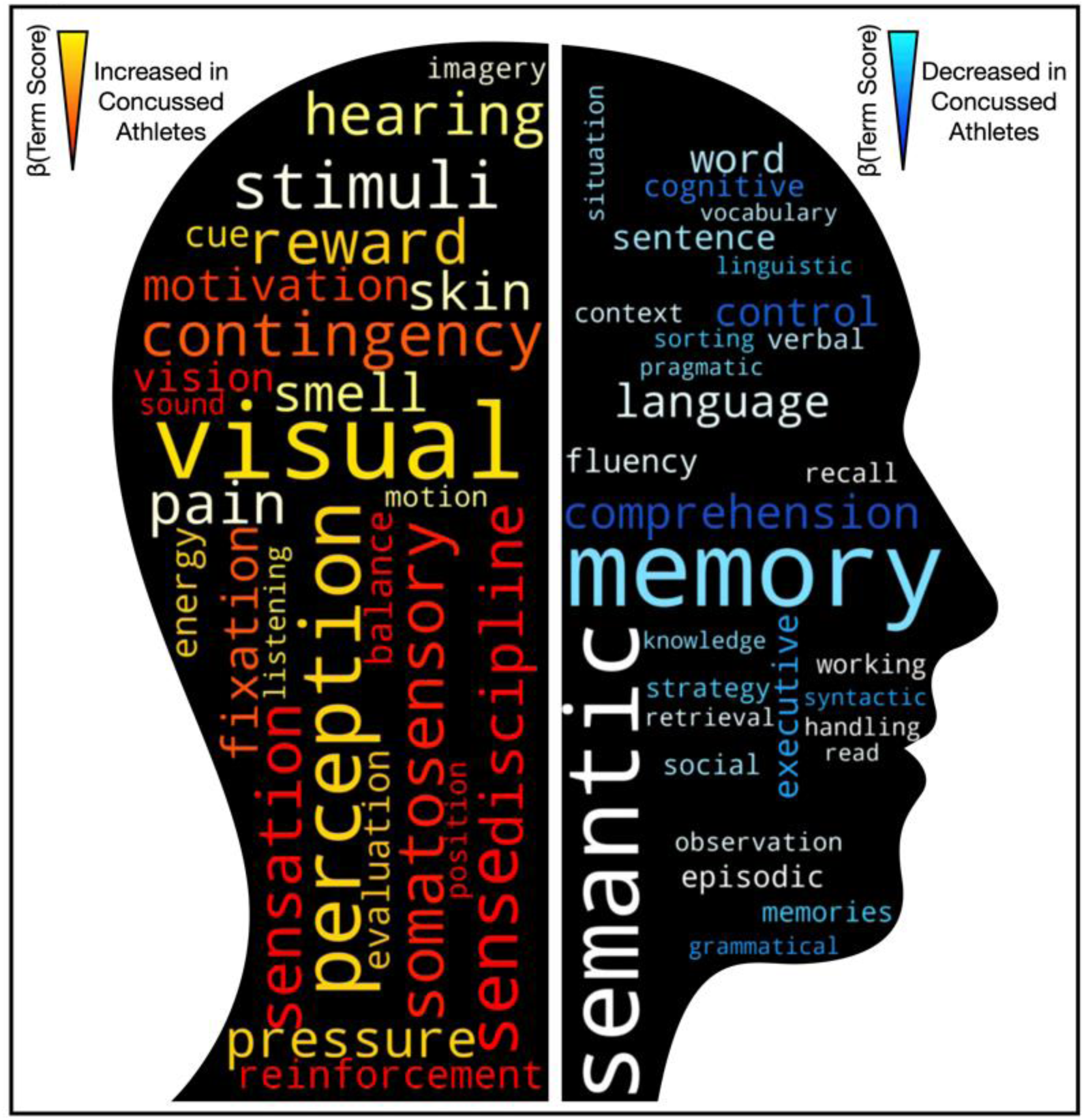
Word-cloud representations of meta-analytic function analysis, derived from 13,459 neuroimaging studies encompassing 5,144 activation pattern terms. Text increasingly red-orange-yellow are those functions putatively enhanced within the athlete group, compared to controls. Reciprocally, text increasingly blue-light blue are those functions putatively attenuated within the athlete group.

## Discussion

### Summary of findings

This study details the structural and functional brain changes of both retired female and retired male athletes with a history of concussion, incurred across a range of sports, compared to matched controls. We reveal an array of brain areas whose structure and function appear to be enhanced within the athlete cohort - across the motor, sensory, coordination, balance, and visual perception domains. We have also demonstrated a reciprocal profile in the concussed cohort, where an array of brain areas exhibiting decreased volume, which when modelled to meta-analytic imaging data, implicates functional attenuation across memory, recall, language, and cognition domains. Separately, with functional MRI we identify a strengthened brain network in athletes, with an enhanced connection between motor and sensory processing regions, where increasing bouts of concussion reverses this effect at key network linkage sites. Figure 5 <u>- below.</u>

The main messages evident from these data suggest that sport plausibly benefits brain structure and function, but that the brains of these individuals may be adversely affected by concussion and mild TBI, and this would attenuate any possible benefit. Attributing these adverse findings to neurodegeneration suggests a relationship between TBI and decreased brain volumes in key regions, most marked in the hippocampus with subsequent compensatory enlargement of the adjacent hippocampal and collateral sulci.

### Promoted structural and functional correlates of sport

We identified that within the athlete cohort, the medial postcentral gyrus, middle orbital gyrus, middle occipital gyrus, transverse temporal gyrus, and lateral orbital gyrus were of increased volume when compared to the controls, which in term meta-analysis related to an increased putative functional enhancement in motor-sensory, perception, coordination, reflexes, and visual domains. Independently then examining the functional connectivity of our cohort similarly demonstrated a functional brain network stronger within the athlete cohort, which characteristically contained functional communities well implicated in motor-sensory and visual processing, with greatest degree counts in nodes of the pre-and post-central gyri, insula, central opercular cortex. Evaluating the eigenvector centrality, broadly a measure of graphical ‘*influence’* of a node within a network[59], confirmed the nodes within the somatomotor, visual and auditory networks to be more *influential* when compared to the remainder. Taken together, these structural and functional findings seem plausible in an athletic group, suggesting motor-sensory functions are raised, both quantified directly with our brain imaging data and that of large-scale meta-analytic repositories.

It is important to note that the enhanced putative brain changes in motor, sensory, balance, visuospatial, and coordination domains in the concussed athletic cohort were still contrasted to a control group who were also physically active (albeit as a leisure activity, rather than at a competitive level). These findings indicate that retired concussed athletes (with a history of participation at a competitive level as an adult) have enhanced brain function in those domains either through practice/repetition and/or through genetic selection for their sport. Furthermore, the concussed athletic volunteers in the study were retired from their sport for 15-40 years, indicative that these individuals therefore must retain these brain enhancements long-term.

### Impacted brain structure and function by repeated concussive head injury across athletes

Whilst we identified a range of brain findings associated with sport, this also revealed an array of reciprocal structural and functional brain changes negatively impacted in the concussed athlete group. At the structural level, we identified the concussed athlete group to demonstrate significantly smaller hippocampi, middle frontal gyri, supramarginal gyri, temporal poles, and inferior frontal gyri compared to the controls, which in meta-analytic term matching related to attenuation across the functional domains of memory, recall, language, and cognition, corroborating and expanding upon smaller scale previous studies using extensive neuropsychological testing[60].

These findings were further amplified in further independent structural analyses of the subcortex, which demonstrated bilateral hippocampal inward deformation in the concussed athlete group. They corroborate and expand upon previous work in smaller-scale studies of similar athletic groups[24, 61]. Importantly, the hippocampal changes were shown to also be directly linked to increasing bouts of concussion. Furthermore, whilst the relationship between hippocampal morphology and age was significantly linked, this affected different margins of the brain region, not least that the relationship between age and hippocampal deformation in both athletes and controls was not significantly different. This critically illuminates that these changes in the hippocampus cannot be merely a function of age but must be driven by other factors distinguishing the groups. To that end, when re-modelling structural brain data but with the supplementation of concussion as an additional covariate, these findings between athletes and controls all became non-significant, supplementing the notion that the history of concussive TBI was the discriminating factor in differential hippocampal volume.

At the functional level, we demonstrated that several links within the connectivity network identified in athletes were weakened with relation to increasing episodes of concussion, which included hemispheric links between the left and right middle frontal gyri, frontal pole, and superior frontal gyri, the links between those paired bilateral regions, as well as the precentral gyrus and insula, superior frontal gyrus and cuneus. Evaluating nodal network influence by eigenvector centrality with regards to concussion revealed a predominance for impact on frontal regions, which seemed to broadly corroborate our structural analyses illustrating volumetrically smaller frontal and temporal regions. Furthermore, we demonstrated that nodal influence between a function of being an athlete with an increasing number of concussions was inversely related: a motor-sensory network uniquely strengthened and influential in the athletes - but weakened with increasing episodes of concussion.

### Implications for cognitive reserve

In identifying an array of brain regions, the volume and function of which are putatively enhanced in the athlete group raises the question of how these findings might implicate cognitive reserve[62]. This point is amplified further given that the athletes were retired from the sports for 15-40 years, therefore clearly retain any functional enhancement either through practice/repetition or through underpinning genetic selection factors for their sport. Multiple previous works suggest that cognitive reserve may be a factor in driving patient recovery from TBI[63-66]. Therefore, future research should explore any relationship between the extent of sporting exposure, cognitive reserve and how that might modify recovery from TBI, and/or offer some protection from adverse neurological consequences.

### Strengths and limitations

Our study design prioritises the recruitment of individuals exposed to repeated episodes of concussion and their long-term follow-up. With that in mind, our group sample sizes, therefore, favoured the exposed group with a smaller number of controls. Notwithstanding that, our controls are closely matched across all other domains quantified. Supplementing the control group size with healthy controls from previous studies seemed less preferable, as otherwise confounds from differently used scanners could have been introduced. Reassurance in our findings comes from the observation that when models comparing cohorts were re-ran with the inclusion of concussion as a variable, all findings became non-significant, suggesting this sporting injury exposure was the main distinguisher in our group. Secondly, our scanning paradigm prioritised the acquisition of high-resolution structural and functional sequences but did not include high-resolution diffusion-weighted sequences. Rather, clinical diffusion sequences were acquired to establish that participants were fit for study inclusion with no major TBI evident. To our strength, we investigated – to our knowledge – the largest sample of its kind across a multi-modal image space engendering brain structure and function. Reassurance also comes with the knowledge that other groups have detailed white-matter-specific diffusion imaging changes to study this knowledge gap[30]. There are a few other potential explanations for these findings. Participants capable of sport may differ neurally from those who are not, producing a difference that is not caused by sport but is a precondition for engaging within it.

## Conclusion

We characterise in detail the structural and functional brain changes apparent in a large sample of retired athletes, who throughout their sporting careers have been exposed to repeated concussive mild TBI. We reveal an array of brain structural and functional changes plausibly related to engagement in sport, linked to putative functional enhancement across the motor, sensory, coordination, balance, and visual processing. These findings illustrate that engagement in such sport may benefit the brain in numerous domains, but also highlights the importance of how it must be protected from the potentially damaging effects of concussive head injury, a notion that may have an impact on cognitive reserve in these individuals. Future work must characterise these brain differences temporally and ascertain if these relate to the development of neurodegenerative disorders.

## Data Availability

All data will be made available to suitably qualified researchers once the study as been completed.

## Abbreviations

fMRI: functional Magnetic Resonance Imaging
NBS: Network-Based Statistics
TBI: Traumatic Brain Injury

## Acknowledgments

The ICHIRF project is currently philanthropically funded by Godolphin Racing, the Injured Jockeys Fund (UK), the Irish Injured Jockeys Fund, the Professional Footballers Association (UK), the National Football League (US), the Concussion Foundation, the Racing Foundation, the British Association of Sport and Exercise Medicine as well as private donations.

The ICHIRF-BRAIN Study would not be possible without the input of the ICHIRF Project Manager, Pippa Theo.

In addition, the authors are indebted to Dr James Ruffle for the statistical analysis, and the invaluable contribution of his colleagues in the Department of Neuroimaging at University College London who modestly declined co-authorship.

## Author Contributions

PM, MT – senior leads for the study, patient recruitment, collaborator recruitment and supervision, and revision of the manuscript for intellectual content.

AB – provided independent review and evaluation of BrainageR data.

RC – provided independent review and evaluation of the research design, analysis of data and outcomes

## Supplementary material

Supplementary Tables 1-3 are available separately on the website -

