## Supplementary Table 1 for "Changes in brain structure and function in a multisport cohort of retired female and male athletes, many years after suffering a concussion. The ICHIRF-BRAIN Study"

| Parameter | Mean | SD | Q2.5 | Q97.5 | 95% CI excl. 0 |
|---|---|---|---|---|---|
| Intercept | 9.47 | 2.68 | 4.34 | 14.88 | * |
| LateralVentricles | -0.33 | 0.22 | -0.77 | 0.11 | |
| ThirdFourthFifthVentricles | -0.32 | 0.22 | -0.76 | 0.11 | |
| NAc | -0.48 | 0.43 | -1.34 | 0.36 | |
| Amyg | -0.28 | 0.42 | -1.11 | 0.53 | |
| Caudate | 0.85 | 0.37 | 0.13 | 1.60 | * |
| Hippocampus | -1.33 | 0.42 | -2.17 | -0.53 | * |
| Pallidum | 0.05 | 0.36 | -0.64 | 0.76 | |
| Putamen | -0.13 | 0.44 | -1.00 | 0.73 | |
| Thalamus | -0.60 | 0.34 | -1.29 | 0.06 | |
| ACgG | 0.02 | 0.35 | -0.67 | 0.71 | |
| Ins | 0.27 | 0.24 | -0.20 | 0.75 | |
| AOrG | 0.44 | 0.45 | -0.43 | 1.32 | |
| AnG | -0.46 | 0.42 | -1.29 | 0.35 | |
| Calc | -0.16 | 0.40 | -0.93 | 0.61 | |
| FO | 0.15 | 0.42 | -0.68 | 0.98 | |
| FRP | -0.39 | 0.31 | -1.00 | 0.21 | |
| FuG | -0.41 | 0.48 | -1.35 | 0.55 | |
| GRe | -0.17 | 0.42 | -1.00 | 0.66 | |
| IOG | -0.82 | 0.47 | -1.75 | 0.08 | |
| ITG | 0.81 | 0.45 | -0.06 | 1.71 | |
| LiG | 0.02 | 0.43 | -0.82 | 0.86 | |
| LOrg | 0.81 | 0.39 | 0.04 | 1.59 | * |
| MCgG | 0.34 | 0.33 | -0.29 | 1.00 | |
| MFC | 0.23 | 0.42 | -0.60 | 1.06 | |
| MFG | -1.36 | 0.45 | -2.25 | -0.50 | * |
| MOG | 0.98 | 0.40 | 0.21 | 1.77 | * |
| MOrG | 0.96 | 0.48 | 0.03 | 1.93 | * |
| MPoG | 0.68 | 0.35 | 0.01 | 1.37 | * |
| MPrG | -0.42 | 0.36 | -1.14 | 0.28 | |
| MSFG | -0.07 | 0.41 | -0.88 | 0.73 | |
| MTG | 0.41 | 0.40 | -0.37 | 1.21 | |
| OCP | -0.59 | 0.38 | -1.36 | 0.14 | |
| OFuG | 0.37 | 0.44 | -0.50 | 1.24 | |
| IFG | -0.69 | 0.25 | -1.20 | -0.22 | * |
| PCgG | 0.62 | 0.41 | -0.18 | 1.44 | |
| PCu | -0.41 | 0.40 | -1.20 | 0.36 | |
| Hi&CollateralSulcus | 2.34 | 0.52 | 1.34 | 3.38 | * |
| PoG | 0.57 | 0.40 | -0.21 | 1.36 | |
| POrG | -0.10 | 0.40 | -0.89 | 0.67 | |

| Parameter | Mean | SD | Q2.5 | Q97.5 | 95% CI excl. 0 |
|---|---|---|---|---|---|
| PrG | 0.27 | 0.41 | -0.51 | 1.08 | |
| SFG | 0.50 | 0.39 | -0.30 | 1.25 | |
| SMC | -0.68 | 0.44 | -1.55 | 0.17 | |
| SMG | -0.96 | 0.36 | -1.69 | -0.27 | * |
| SOG | 0.67 | 0.40 | -0.12 | 1.46 | |
| SPL | -0.64 | 0.33 | -1.29 | 0.01 | |
| STG | 0.28 | 0.42 | -0.54 | 1.10 | |
| TMP | -0.91 | 0.38 | -1.67 | -0.17 | * |
| TTG | 0.82 | 0.34 | 0.17 | 1.49 | * |
| $Age^2$ | 0.00 | 0.00 | 0.00 | 0.00 | |
| Gender | -0.35 | 0.79 | -1.91 | 1.20 | |

**Supplementary Table 1:** Results of the Bayesian logit model of brain volume differences between athletes and controls. Table depicts mean posterior estimate, standard deviation (SD), 95% credible interval, with asterisk demonstrating if the credible interval of the posterior estimate did not cross zero and hence was significant. Athletes are treated as the positive class with positive posterior estimates signifying increased volumes in the athletes, and reciprocally negative values signifying decreased volumes. Abbreviations: Amyg., amygdala; ACgG, anterior cingulate gyrus; AOrG, anterior orbital gyrus; AnG, angular gyrus; Calc, calcarine cortex; FO, frontal operculum; FRP, frontal pole; FuG, occipital fusiform gyrus; GRe, gyrus rectus; Hi, hippocampal [sulcus]; Ins, insula; IFG, inferior frontal gyrus; IOG, inferior occipital gyrus; ITG, inferior temporal gyrus; LiG, lingual gyrus; LOrg, lateral orbital gyrus; MCgG, middle cingulate gyrus; MFC, medial frontal cortex; MFG, middle frontal gyrus; MOG, middle occipital gyrus; MOrG, medial orbital gyrus; MPoG, postcentral gyrus medial segment; MPrG, precentral gyrus medial segment; MSFG, superior frontal gyrus medial segment; MTG, middle temporal gyrus; NAc, nucleus accumbens; OCP, occipital pole; OFuG, occipital fusiform gyrus; PCgG, posterior cingulate gyrus; PCu, precuneus; PoG, postcentral gyrus; POrG, posterior orbital gyrus; PrG, precentral gyrus; SFG, superior frontal gyrus; SMC, supplementary motor cortex; SMG, supramarginal gyrus; SOG, superior occipital gyrus; SPL, superior parietal lobule; STG, superior temporal gyrus; TMP, temporal pole; TTG, transverse temporal gyrus.
