## Supplementary Table 3 for "Changes in brain structure and function in a multisport cohort of retired female and male athletes, many years after suffering a concussion. The ICHIRF-BRAIN Study"

**Bayesian Poisson ridge regression**

Number of obs = 107
Number of vars = 99
MCMC Samples = 100000
MCMC Burnin = 50000
MCMC Thinning = 5
Overdispersion = 1.002
Pseudo R2 = 0.5794
WAIC = 364.48

| **Parameter** | **mean(Coef)** | **std(Coef)** | **[95% Cred.** | **Interval]** | **tStat** | **Rank** | | **ESS** |
|---|---|---|---|---|---|---|---|---|
| GenderM:SPL | -0.54935 | 0.10344 | -0.75994 | -0.35867 | -5.311 | 1 | ** | 121 |
| GenderM:PCu | -0.30912 | 0.08787 | -0.47891 | -0.13037 | -3.518 | 2 | ** | 133 |
| GenderM:FuG | -0.34664 | 0.10907 | -0.56355 | -0.12721 | -3.178 | 3 | ** | 96 |
| GenderM:PoG | 0.34854 | 0.10219 | 0.13614 | 0.54132 | 3.411 | 4 | ** | 138 |
| GenderM:ThirdFourthFifthVentricles | 0.29229 | 0.06895 | 0.15191 | 0.42356 | 4.239 | 5 | ** | 169 |
| GenderM | 0.86718 | 0.32083 | 0.24796 | 1.48399 | 2.703 | 6 | ** | 90 |
| GenderM:AnG | -0.29519 | 0.09349 | -0.48323 | -0.12117 | -3.157 | 6 | ** | 114 |
| GenderM:OCP | -0.25019 | 0.08077 | -0.42035 | -0.10292 | -3.098 | 8 | ** | 122 |
| GenderM:NAc | 0.23363 | 0.09211 | 0.05485 | 0.41981 | 2.537 | 9 | ** | 89 |
| MOG | 0.20917 | 0.07573 | 0.07705 | 0.36814 | 2.762 | 10 | ** | 115 |
| GenderM:LateralVentricles | -0.12812 | 0.05049 | -0.23179 | -0.0306 | -2.538 | 11 | ** | 131 |
| PCu | 0.1662 | 0.0744 | 0.02184 | 0.31664 | 2.234 | 12 | ** | 116 |
| GenderM:MFG | -0.21585 | 0.09504 | -0.39534 | -0.02829 | -2.271 | 12 | ** | 121 |
| GenderM:SMG | 0.21901 | 0.08282 | 0.06435 | 0.38767 | 2.644 | 12 | ** | 112 |
| MTG | -0.16217 | 0.07417 | -0.31838 | -0.0217 | -2.186 | 15 | ** | 101 |
| GenderM:MOG | -0.23986 | 0.10887 | -0.46836 | -0.04586 | -2.203 | 15 | ** | 106 |
| GenderM:OFuG | -0.23122 | 0.0828 | -0.40446 | -0.07877 | -2.793 | 15 | ** | 247 |
| ITG | 0.16009 | 0.08023 | 0.01061 | 0.32345 | 1.995 | 18 | ** | 110 |
| PrG | 0.14965 | 0.06299 | 0.02884 | 0.27637 | 2.376 | 18 | ** | 138 |
| GenderM:Caudate | 0.1811 | 0.07965 | 0.02596 | 0.33945 | 2.274 | 20 | ** | 103 |

| | | | | | | | | |
|---|---|---|---|---|---|---|---|---|
| GenderM:IOG | 0.20715 | 0.10121 | 0.01069 | 0.42128 | 2.047 | 21 | ** | 182 |
| MSFG | -0.15211 | 0.08443 | -0.32986 | 0.00461 | -1.802 | 22 | * | 78 |
| GenderM:Ins | -0.09982 | 0.05097 | -0.20722 | -0.00363 | -1.958 | 22 | ** | 108 |
| GenderM:MOrG | 0.20013 | 0.10588 | 0.00566 | 0.41811 | 1.89 | 24 | ** | 120 |
| GenderM:MTG | 0.1592 | 0.087 | -0.01057 | 0.33409 | 1.83 | 24 | * | 78 |
| GenderM:Age | -0.00013 | 0.00008 | -0.00029 | 0.00002 | -1.626 | 26 | * | 82 |
| `Hi&CollateralSulcus` | 0.12778 | 0.07307 | -0.01445 | 0.26483 | 1.749 | 27 | * | 169 |
| POrG | -0.11941 | 0.06967 | -0.25617 | 0.01993 | -1.714 | 27 | * | 132 |
| GenderM:Pallidum | 0.14195 | 0.08836 | -0.03598 | 0.31653 | 1.607 | 29 | * | 144 |
| SMG | -0.11382 | 0.06074 | -0.23125 | 0.00641 | -1.874 | 30 | * | 124 |
| FRP | 0.106 | 0.06293 | -0.01945 | 0.22465 | 1.684 | 31 | * | 114 |
| MPrG | -0.10365 | 0.06561 | -0.23161 | 0.03413 | -1.58 | 32 | * | 105 |
| GenderM:GRe | -0.15773 | 0.09273 | -0.33986 | 0.01851 | -1.701 | 32 | * | 98 |
| Putamen | 0.10102 | 0.06966 | -0.03465 | 0.2428 | 1.45 | 34 | * | 138 |
| Calc | -0.09092 | 0.05624 | -0.20519 | 0.01388 | -1.617 | 35 | * | 126 |
| IOG | 0.11541 | 0.08487 | -0.05697 | 0.28564 | 1.36 | 35 | * | 125 |
| LOrg | -0.11118 | 0.07688 | -0.2672 | 0.03352 | -1.446 | 35 | * | 120 |
| GenderM:Calc | -0.12133 | 0.07306 | -0.26179 | 0.02538 | -1.661 | 35 | * | 99 |
| LiG | 0.09827 | 0.07304 | -0.0382 | 0.24683 | 1.345 | 39 | * | 142 |
| GenderM:Thalamus | -0.10675 | 0.07921 | -0.26684 | 0.04711 | -1.348 | 39 | * | 187 |
| GenderM:AOrG | 0.13187 | 0.08743 | -0.04022 | 0.30761 | 1.508 | 39 | * | 166 |
| PoG | -0.0871 | 0.06135 | -0.20941 | 0.03174 | -1.42 | 42 | * | 149 |
| IFG | -0.0389 | 0.0319 | -0.10292 | 0.02229 | -1.219 | 43 | * | 136 |
| GenderM:SOG | 0.10175 | 0.0869 | -0.07282 | 0.27354 | 1.171 | 43 | * | 97 |
| MPoG | -0.0912 | 0.06228 | -0.21248 | 0.0316 | -1.464 | 45 | * | 153 |
| GenderM:ITG | -0.1073 | 0.10054 | -0.30292 | 0.08973 | -1.067 | 45 | | 126 |
| GenderM:IFG | 0.04514 | 0.04573 | -0.04163 | 0.13971 | 0.987 | 45 | | 118 |
| GenderM:MFC | 0.11051 | 0.09312 | -0.07185 | 0.29488 | 1.187 | 48 | * | 109 |
| Pallidum | -0.0735 | 0.06828 | -0.20149 | 0.05819 | -1.076 | 49 | | 137 |
| FO | 0.0845 | 0.07962 | -0.06574 | 0.23823 | 1.061 | 49 | | 115 |

| | | | | | | | |
|---|---|---|---|---|---|---|---|
| GenderM:Amyg | 0.07444 | 0.08618 | -0.10181 | 0.24073 | 0.864 | 49 | 116 |
| GenderM:FRP | -0.07646 | 0.07824 | -0.22139 | 0.08079 | -0.977 | 49 | 129 |
| NAc | -0.06905 | 0.07479 | -0.21506 | 0.0821 | -0.923 | 53 | 115 |
| TTG | -0.07111 | 0.06382 | -0.20137 | 0.05778 | -1.114 | 54 | 161 |
| GenderM:LiG | 0.09507 | 0.10239 | -0.11493 | 0.29612 | 0.929 | 54 | 123 |
| AOrG | 0.07058 | 0.06275 | -0.05154 | 0.1925 | 1.125 | 56 | 138 |
| GenderM:MPrG | 0.08677 | 0.08373 | -0.08377 | 0.24846 | 1.036 | 57 | 149 |
| FuG | -0.05552 | 0.07944 | -0.20643 | 0.09938 | -0.699 | 58 | 113 |
| GenderM:LOrg | 0.08812 | 0.08741 | -0.07881 | 0.27334 | 1.008 | 58 | 120 |
| GenderM:MSFG | -0.02429 | 0.09835 | -0.20053 | 0.17438 | -0.247 | 60 | 86 |
| GenderM:PrG | 0.07417 | 0.08947 | -0.09942 | 0.24949 | 0.829 | 60 | 157 |
| Hippocampus | 0.03976 | 0.06844 | -0.09228 | 0.1779 | 0.581 | 62 | 139 |
| OCP | -0.05431 | 0.05647 | -0.16383 | 0.05676 | -0.962 | 62 | 173 |
| PCgG | 0.03872 | 0.07815 | -0.1061 | 0.19948 | 0.495 | 62 | 93 |
| GenderM:Hippocampus | -0.05326 | 0.08324 | -0.2194 | 0.10486 | -0.64 | 62 | 111 |
| GenderM:ACgG | -0.03352 | 0.10372 | -0.23967 | 0.16418 | -0.323 | 62 | 132 |
| GenderM:PCgG | -0.02865 | 0.10088 | -0.2293 | 0.1583 | -0.284 | 62 | 83 |
| GenderM:`Hi&CollateralSulcus` | -0.01285 | 0.1061 | -0.22333 | 0.18816 | -0.121 | 62 | 129 |
| GenderM:TMP | 0.07327 | 0.07947 | -0.08302 | 0.22898 | 0.922 | 62 | 149 |
| Amyg | 0.02326 | 0.07656 | -0.11938 | 0.17793 | 0.304 | 70 | 135 |
| ACgG | -0.04018 | 0.0706 | -0.17553 | 0.09513 | -0.569 | 70 | 58 |
| Ins | -0.00165 | 0.04214 | -0.07301 | 0.08909 | -0.039 | 70 | 90 |
| GRe | 0.04119 | 0.06881 | -0.09698 | 0.1731 | 0.599 | 70 | 61 |
| MFC | -0.02277 | 0.07316 | -0.16475 | 0.11756 | -0.311 | 70 | 99 |
| SFG | 0.01634 | 0.07412 | -0.1268 | 0.16155 | 0.22 | 70 | 152 |
| SMC | -0.02994 | 0.06903 | -0.16205 | 0.10126 | -0.434 | 70 | 141 |
| SPL | 0.04731 | 0.06844 | -0.08598 | 0.18453 | 0.691 | 70 | 119 |
| Age | -0.0001 | 0.0001 | -0.00028 | 0.00009 | -1.028 | 70 | 125 |
| GenderM:Putamen | 0.04054 | 0.08906 | -0.12721 | 0.21779 | 0.455 | 70 | 120 |
| Thalamus | 0.0383 | 0.06077 | -0.08064 | 0.15408 | 0.63 | 80 | 152 |

| | | | | | | | |
|---|---|---|---|---|---|---|---|
| AnG | 0.03677 | 0.06684 | -0.09579 | 0.16593 | 0.55 | 80 | 115 |
| MFG | 0.0164 | 0.07028 | -0.12069 | 0.15193 | 0.233 | 80 | 146 |
| GenderM:SFG | -0.00387 | 0.08023 | -0.17415 | 0.15095 | -0.048 | 80 | 154 |
| GenderM:SMC | 0.02524 | 0.09244 | -0.15552 | 0.20031 | 0.273 | 80 | 122 |
| GenderM:STG | -0.02581 | 0.085 | -0.18713 | 0.14258 | -0.304 | 80 | 154 |
| GenderM:TTG | -0.06652 | 0.08112 | -0.22672 | 0.09109 | -0.82 | 80 | 165 |
| MOrG | 0.00372 | 0.07352 | -0.13282 | 0.15275 | 0.051 | 87 | 118 |
| SOG | -0.02788 | 0.06769 | -0.16184 | 0.11139 | -0.412 | 87 | 129 |
| STG | 0.02253 | 0.06721 | -0.11202 | 0.1532 | 0.335 | 87 | 167 |
| GenderM:FO | -0.02305 | 0.09564 | -0.2165 | 0.16556 | -0.241 | 87 | 127 |
| GenderM:POrG | 0.03546 | 0.09153 | -0.15062 | 0.20991 | 0.387 | 87 | 120 |
| ThirdFourthFifthVentricles | -0.02874 | 0.04856 | -0.12633 | 0.06173 | -0.592 | 92 | 191 |
| Caudate | -0.00859 | 0.06599 | -0.13942 | 0.12067 | -0.13 | 92 | 103 |
| MCgG | -0.01552 | 0.07282 | -0.16653 | 0.12426 | -0.213 | 92 | 104 |
| OFuG | -0.0099 | 0.0664 | -0.13429 | 0.12467 | -0.149 | 92 | 170 |
| TMP | -0.01292 | 0.06914 | -0.14853 | 0.12729 | -0.187 | 92 | 128 |
| GenderM:MCgG | 0.03545 | 0.09043 | -0.14658 | 0.21145 | 0.392 | 92 | 103 |
| LateralVentricles | -0.01007 | 0.03643 | -0.07881 | 0.06709 | -0.276 | 98 | 126 |
| GenderM:MPoG | -0.01439 | 0.08619 | -0.18483 | 0.15883 | -0.167 | 98 | 157 |
| _cons | 2.19773 | 0.36136 | 1.48615 | 2.90957 | . | . | . |

**Supplementary Table 3:** Results of the Bayesian regression model of concussion as a function of gender interacting with brain volumetric changes. Table depicts mean posterior estimate, standard deviation (SD), 95% credible interval, with asterisk demonstrating if the credible interval of the posterior estimate did not cross zero and hence was significant. Significant negative posterior estimates relate to decreasing volumetry related to that given interaction, in accordance with increasing episodes of concussion. For example, row one depicts an interaction between males and decreased volume to the superior parietal lobule, for an increase in the number of episodes of concussion. Factors are ranked by their predictive fidelity. Abbreviations: Amyg., amygdala; ACgG, anterior cingulate gyrus; AOrG, anterior orbital gyrus; AnG, angular gyrus; Calc, calcarine cortex; FO, frontal operculum; FRP, frontal pole; FuG, occipital fusiform gyrus; GRe, gyrus rectus; Hi, hippocampal [sulcus]; Ins, insula; IFG, inferior frontal gyrus; IOG, inferior occipital gyrus; ITG, inferior temporal gyrus; LiG, lingual gyrus; LOrg, lateral orbital gyrus; MCgG, middle cingulate gyrus; MFC, medial frontal cortex; MFG, middle frontal gyrus; MOG, middle occipital gyrus; MOrG, medial orbital gyrus; MPoG, postcentral gyrus medial segment; MPrG, precentral gyrus medial segment; MSFG, superior frontal gyrus medial segment; MTG, middle temporal gyrus; NAc, nucleus accumbens; OCP, occipital pole; OFuG, occipital fusiform gyrus; PCgG, posterior cingulate gyrus; PCu, precuneus; PoG, postcentral gyrus; POrG, posterior orbital gyrus; PrG, precentral gyrus; SFG, superior frontal gyrus; SMC, supplementary motor cortex; SMG, supramarginal gyrus; SOG, superior occipital gyrus; SPL, superior parietal lobule; STG, superior temporal gyrus; TMP, temporal pole; TTG, transverse temporal gyrus.
